# Factors Associated with Coronavirus (COVID-19) Deaths and Infections: A Cross Country Evidence

**DOI:** 10.1101/2020.11.02.20183236

**Authors:** Shafiun Nahin Shimul, Fariha Kadir, Muhammad Ihsan- Ul- Kabir

## Abstract

Though most of the countries across the world are crippled with COVID-19, there has been substantial variations in death and infection rates. While some countries are overwhelmed, a few are spared. Little is known what explains this variation. This study attempts to understand the covariates of death and infection rates of COVID-19 across countries using multivariate regression analysis and least absolute shrinkage and selection operator (LASSO) regression. The OLS estimates show that the aging population and hospital bed per capita are significantly associated with the fatality rate of COVID-19, while urbanization has a positive correlation with the inflection rate. The study suggests that an increase in health systems capacity can significantly reduce the fatality rates due to COVID-19.

## 1. Introduction

The outbreak of coronavirus disease 2019 (COVID-19) caused by the novel coronavirus, first reported on 31^st^ December 2019, in Wuhan, China that has since spread to 215 countries and regions globally. As of 23^rd^ June 2020, there were more than nine million confirmed cases (n=8,985,016) and 472,216 deaths^1^. According to World Health Organization, preliminary *R*_*o*_ estimate of COVID-19 is reported as 1.4-2.5 (WHO, 2020) indicating its larger infectiousness. However, while it has walloped many countries, a great number of countries has got spared. In addition, there has been significant variations in deaths across countries. Data from China, South Korea, Italy, and Iran suggest that the case fatality rate (CFR) increases drastically with age and is higher in people with COVID-19 and underlying comorbidities (The Novel Coronavirus Pneumonia Emergency Response Epidemiology Team, 2020). Targeted social distancing for these groups could be the most effective way to reduce morbidity and concurrent mortality. Advanced in age and having chronic conditions, such as obesity, diabetes, respiratory disease, kidney disease and cardiovascular diseases (CVD) have been linked with more severe COVID-19 symptoms, often leading to the development of acute respiratory distress syndrome (ARDS) and progression from ARDS to death (ChaominWu, et al., 2020)

Another study finds that person’s occupation may expose them to risk by the nature of their job. Work involving constant human contact, interaction with others or caring for people means that risk of infection spread through droplets is higher (Rule, et al., 2018). In response to COVID-19, occupation is likely to be a direct determinant of infection and an indirect determinant of disease severity and mortality through the relationship between occupational social class and comorbidities (Koh, 2020).

Low income and lower education level are indirectly associated with a number of factors that may increase the risk of developing severe forms of COVID-19, such as increased prevalence of smoking, poor nutrition, and housing conditions which could suppress the immune system. A recent systematic review of five retrospective or prospective studies reported that smoking is most likely associated with adverse outcomes of COVID-19 (Vardavas & Nikitara, 2020). In order to define the grounds which are most likely to be associated with COVID-19 outbreaks, high-quality data on socioeconomic factors are urgently needed, which will have important implications in the development of health system measures. (Khalatbari-Soltani, Cumming, & Delpierre, 2020).

Even though the COVID-19 has spread across the globe, not all countries got equally affected. While the spread ballooned in some countries, some countries got relatively spared. Non-pharmaceutical interventions have played a significant role. Still, a lot of unknowns are there as well. Even though from the various sources, it was seen that ageing population are vulnerable, health system’s capacity can also dramatically reduce that vulnerably. While a lot of micro or patient level data and study are emerging, and can better answer the underlying factors of case fatality rates and infection rates once COVID-19 reaches in its ending phase, a cross-country comparison can elicit some demographic and health system covariates of the death and infection rates. A few such effort, till to date, however, has been observed. In this study, we attempt to evaluate the extent of underlying factors with the latest available data using various empirical estimation methods.

## 2. Related Literature

We identify published studies through a rapid review and observe that myriads of social and economic requisites have been attributed as potential determinants for the empirical variety in the coronavirus outcome. Hardly any of determinants, however, are able to robustly unravel the extent of the coronavirus pandemic. Stojkoski, Utkovski, Jolakoski, Tevdovski, & Kocarev (2020) recommend that population size and government health expenditure are the two determinants strongly related to the coronavirus cases. Their study finds out that more populated economies show greater resistance to being infected by the virus, whereas countries with larger government expenditure display greater susceptibility to the virus infection.

Rodrigues, Prata, & Camargo (2020) investigate the regional differences in the occurrence of COVID-19 in Brazil and its relationship with climatic and demographic factors by using a polynomial regression model in terms of the number of days of transmission, demographic density, city population and climatic factors. Their results show that temperature variation maintains a relationship with the reduction in the number of COVID-19 cases. Depending on the contagion process in progress, the expected reduction in the number of cases of COVID- 19 is −3.4% for each increase of 1°C.

Qiu, Chen, & Shi (2020) examines the transmission dynamics of the novel coronavirus 2019, considering both within- and between-city transmissions in China. Using a machine learning method (Lasso), their estimates show that the actual population flow from the outbreak source poses a higher risk to the destination than other factors such as geographic proximity and similarity in economic conditions, and there is evidence that people’s responses can break the chain of infections. Changes in weather conditions also induce exogenous variations in past infection rates, which allows to identify the causal impact of past infections on new cases.

A systematic review conducted by Khalatbari-Soltani, Cumming, & Delpierre (2020) shows that the impact of socioeconomic determinants on the COVID-19 outbreak fluctuate in high-income-countries over low-and middle-income countries (LMICs). According to this study, the community spread of disease COVID-19 appearing initially from abroad, will depend upon the particular local infrastructures and social inequalities in each context. For instance, the usual public health practices of social distancing including lockdowns, quarantine and self-isolation are hardly performed in LMICs. Additionally, such income settings where absolute poverty is a pivotal concern, access to basic needs like food, housing, water and sanitation will have an immense distinction on how easily people can practice any social distancing measures to isolate older adults or vulnerable people. What is more, in many parts of the world, health is not free on the verge of need and health care systems, which are outstretched at the best of times, will become quickly inundated.

Containing contact rates is a vital strategy for outbreak control which also hinges on population densities. Keeping to more than 1 meter distance between people coughing and sneezing, as recommended by the WHO becomes more intricate with higher population densities (WHO, 2020).

Henceforth, avoiding circumstances with higher population densities will be a necessary requirement to limit the spread of COVID-19 (Rocklöv & Sjödin, 2020).

A study reveals that social distancing measures reduce the epidemic value of the effective reproduction number *R*_0_ by about 60% or less, if the intrinsic transmission potential declines in the warm summer months (Anderson, Heesterbeek, & Klinkenberg, 2020). For good measure, broader-scale social distancing provides time for the health systems to treat cases and increase capacity for treatments to be developed in the longer term. In the same manner, experts agree that there is no determinant strongly related to the coronavirus deaths per million population.

A major contribution of the present paper is a stepping stone towards more comprehensive understanding of the relationship between infections as well as deaths of COVID-19 with socio-economic or macro-economic determinants.

## 3. Data and Estimation Method

### Data

Two major data sources are used in this study: *Our World in Data COVID-19 Dataset*^2^ by the University of Oxford, and the World Banks’ *World Development Indicators* (WDI^3^). In our empirical estimations, we use values of population ages 70 years and above (% of total population); population density; urban population (% of total population); hospital bed per 1,000 population; immunization, measles (% of children ages 12-23 months); smoking prevalence, total (ages 15+). We used the mean values of these variables for the period of 2008-2019. Values of case fatality rate (CFR); total tests per thousand; total cases per million; cardiovascular diseases (CVD) death rate; and extreme poverty rate were collected from *Our World in Data COVID-19 Dataset* by the University of Oxford. Values for tropical country dummy (=1 for tropical countries) were collected from *World Population Review*^4^. Our cross-section data set consists of observations for each of the 186 countries of the world. The data is up to June 13, 2020. Figures in the appendix provide some relationship between/among the variables used in this study.

### Estimation method

In our empirical estimations, we use *t*-test, correlation matrix, and Least Absolute Shrinkage and Selection Operator (LASSO) to find out the correlation of the determinants of COVID-19. We draw a test on the equality of means by performing *t*-test. We employ the difference in means of seven factor variables. Taking into account ageing population (70 years and above) of 10%, the variable is valued 1 if a country has more than or equal 10% of its population aged 70 years and above, and 0 otherwise. Similarly, we use the other six factor variables including 3.5 hospital beds per 1,000 population, 80% urban population, tropical country, CVD death rate of 300, 80% measles immunization, and smoking prevalence of 25%. We also report pairwise correlation coefficients between the variables. The choice of cut-point point is the mean/median values, unless otherwise stipulated.

#### OLS & LASSO

While t-test and correlation test can provide some insights on the relationships between variables, they cannot separate out the effects of confounding variables, and therefore, the conclusion made from these could be spurious. Regression analysis can solve this problem of omission of control variables. To understand the effects of demographic, health system and other correlates of death rates and infection, we use the following regression function:

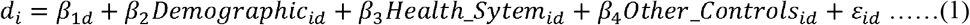

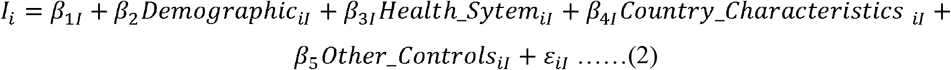

Here, *d* is death rates, *I* is the infection rate per million population and in the right hand side we have all relevant determinants.

First, we applied Ordinary Least Square (OLS) regression to understand the determinants of both infection and death rates. However, it is extremely likely that right hand co-variates are correlated. And if the determining variables are correlated, we might not be able to precisely estimates the coefficients resulting non-rejection of null-hypothesis, i.e. inference will be plagued. Another issue, since COVID-19 is still unknown to us, and so a clear modeling framework is not possible. LASSO have been extremely popular among the data scientists for prediction and model selection. We also applied LASSO—a machine learning tool that accept some level of bias to minimize the variance and select an appropriate model from the data. Though traditional LASSO measures are not meant for inference rather they are better suited for predictions, with some medication in the process we can apply LASSO for inference. We applied double-selection lasso linear regression model for inference. In this process, instead of running a single regression, the following process is followed:

Step 1: running LASSO of key variables on other controls

Step 2: running LASSO of left-hand side variable on other controls

Step 3: finding the union of the selected variables from both step 1 and step 2 and estimate predicated value

Step 3: regressing left hand side variable on key variables and predicated value from step 3

Step 5: making inference

For details of this methods see (Belloni, 2016) & (StataCorp., 2019)

## 4. Findings and Discussion

In this section, we report the descriptive statistics in Table 1, and estimates of correlation matrix, *t*-test, and regression in later tables. For more relations among variables see Figure 1-Figure 6 of appendix.

**Table 1:** Descriptive Statistics.

| Variables | Obs | Mean | Std. Dev. | Min | Max |
| --- | --- | --- | --- | --- | --- |
| Case fatality rate (CFR) | 186 | 3.55 | 3.9 | 0 | 21.99 |
| Total tests per thousand | 83 | 38.62 | 44.56 | 0.44 | 241.45 |
| Total cases per million | 186 | 1,470.81 | 2,753.66 | 0.89 | 26,583.27 |
| Population ages 70 years and above (% of total population) | 173 | 5.60 | 4.27 | 0.53 | 18.49 |
| Tropical country dummy (=1 for tropical countries) | 166 | 0.392 | 0.49 | 0 | 1 |
| Population density | 185 | 265.28 | 840.40 | 0.14 | 7,915.73 |
| Urban population (% of total population) | 186 | 59.9 | 23.27 | 11.52 | 100 |
| Cardiovascular diseases (CVD) death rate | 175 | 256.26 | 118.64 | 79.37 | 724.42 |
| Hospital bed per 1,000 population | 158 | 3.03 | 2.41 | 0.2 | 13.63 |
| Immunization, measles (% of children ages 12-23 months) | 170 | 87.74 | 12.45 | 40.18 | 99 |
| Smoking prevalence, total (ages 15+) | 138 | 22.39 | 9.67 | 2.27 | 47.03 |
| Extreme poverty rate | 117 | 13.71 | 20.67 | 0.1 | 77.6 |

**Figure 1:**
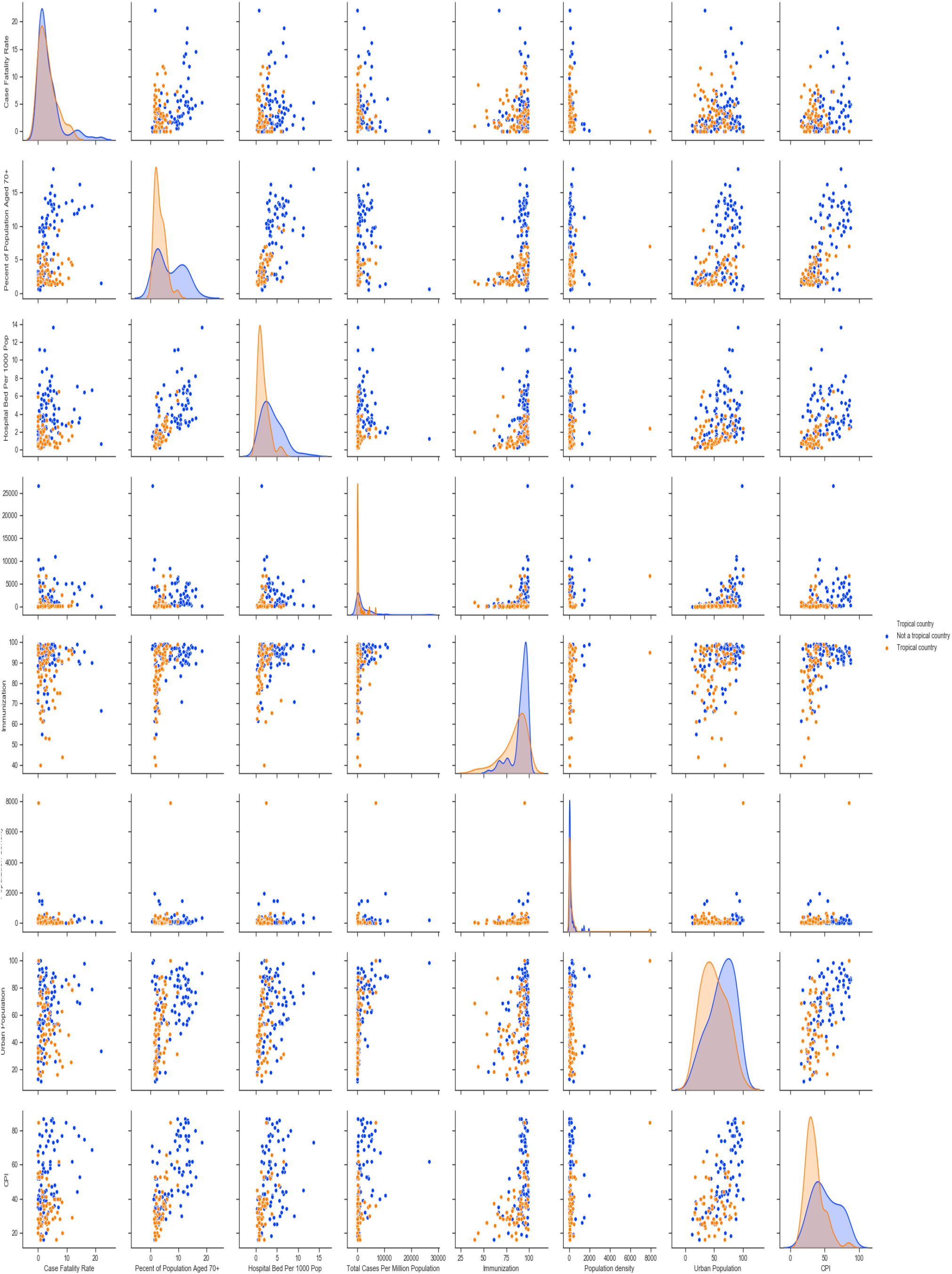
Correlation among variables

**Figure 2:**
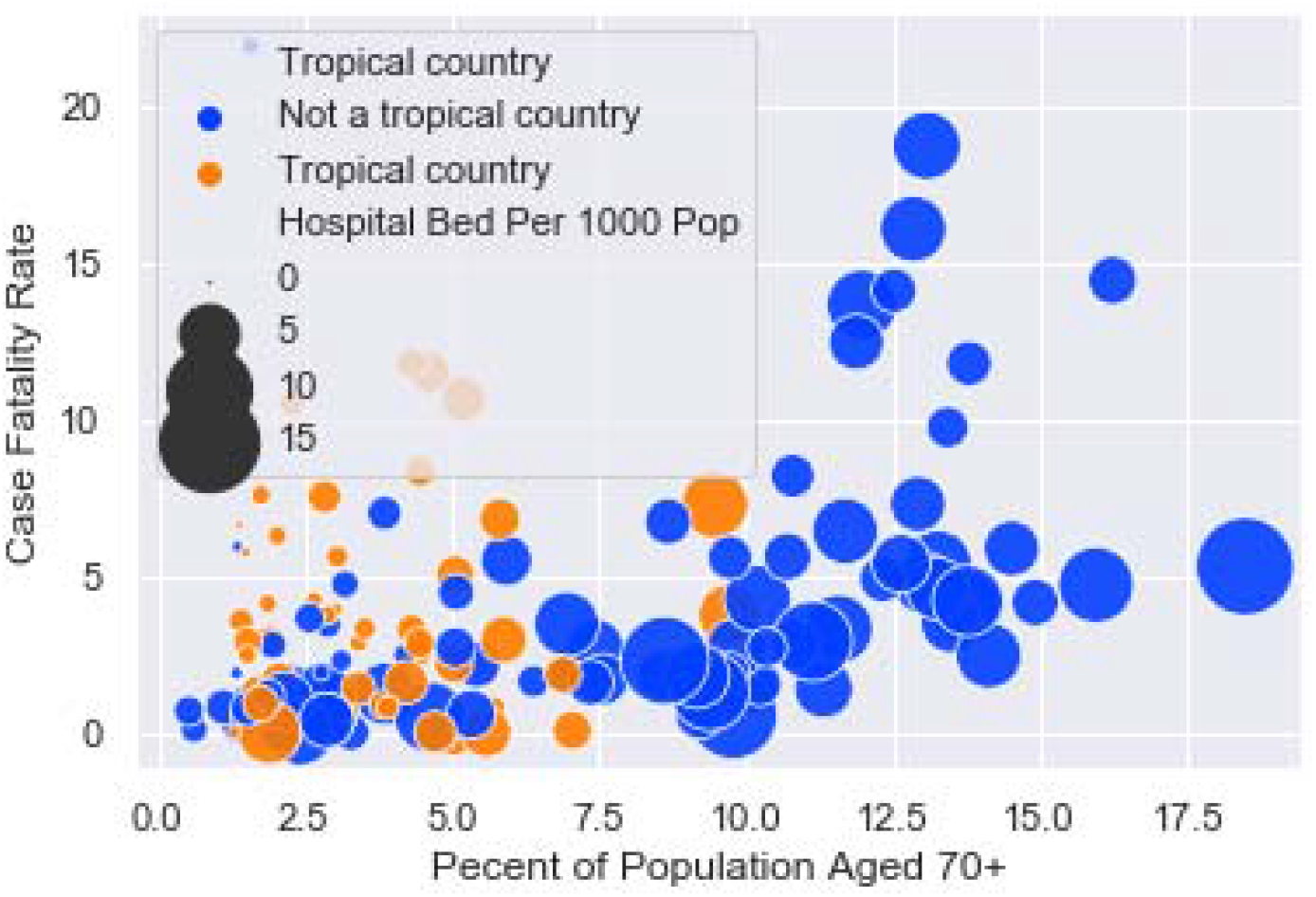
Ageing and case fatality by hospital beds

**Figure 3:**
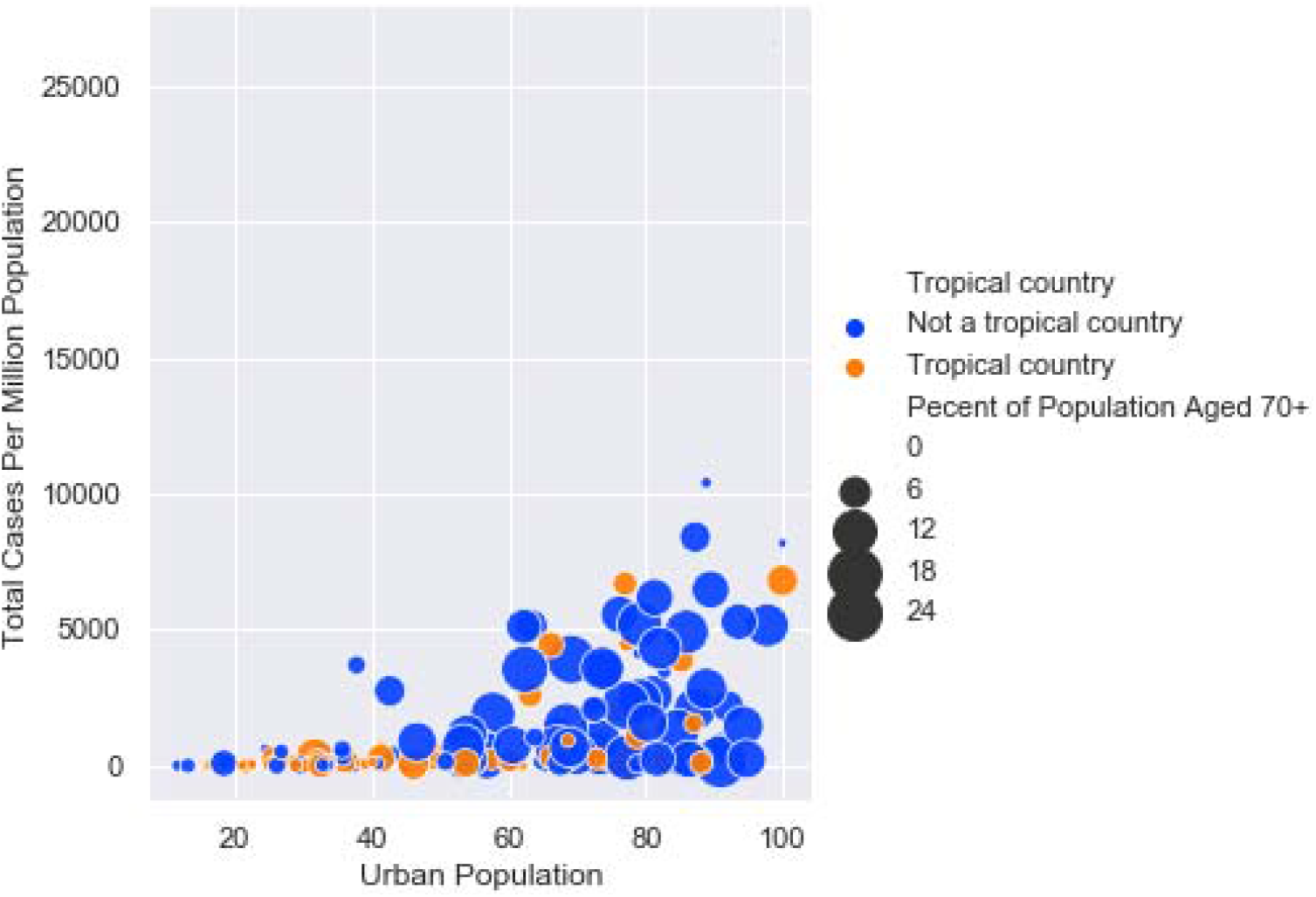
Urban population, infection by ageing population

**Figure 4:**
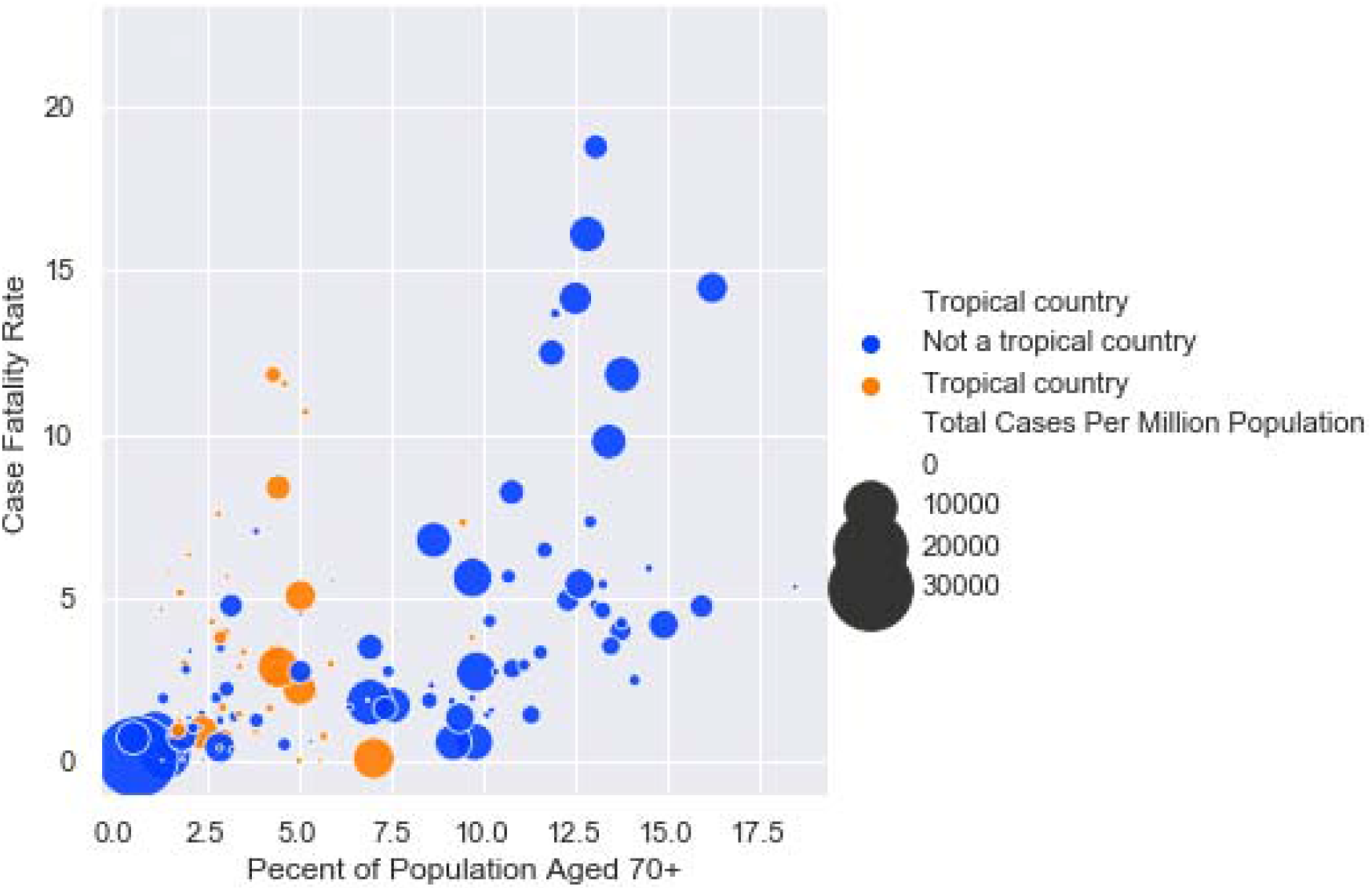
Ageing and case-fatality rate by tests

**Figure 5:**
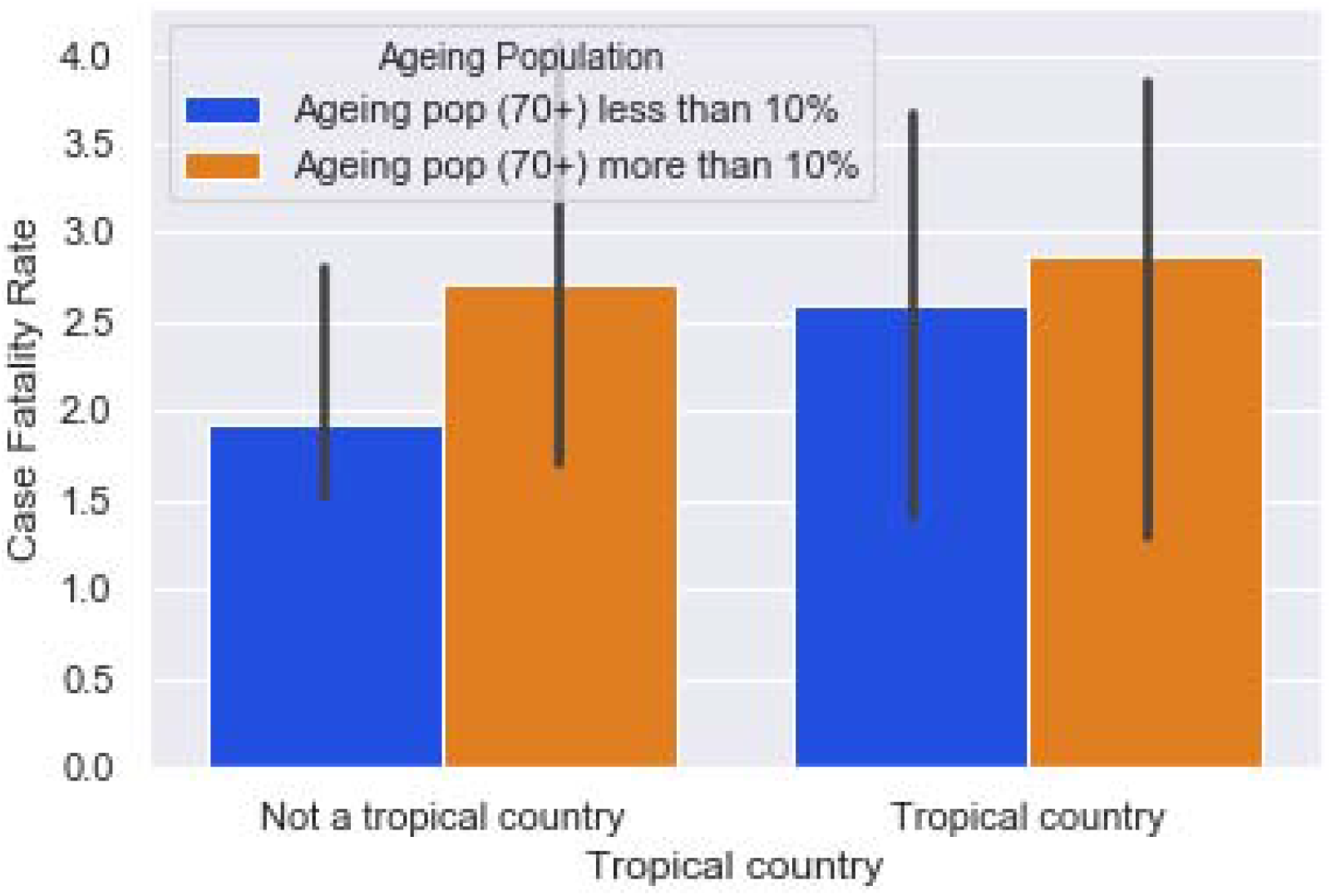
Case fatality rate by tropical status of the country

**Figure 6:**
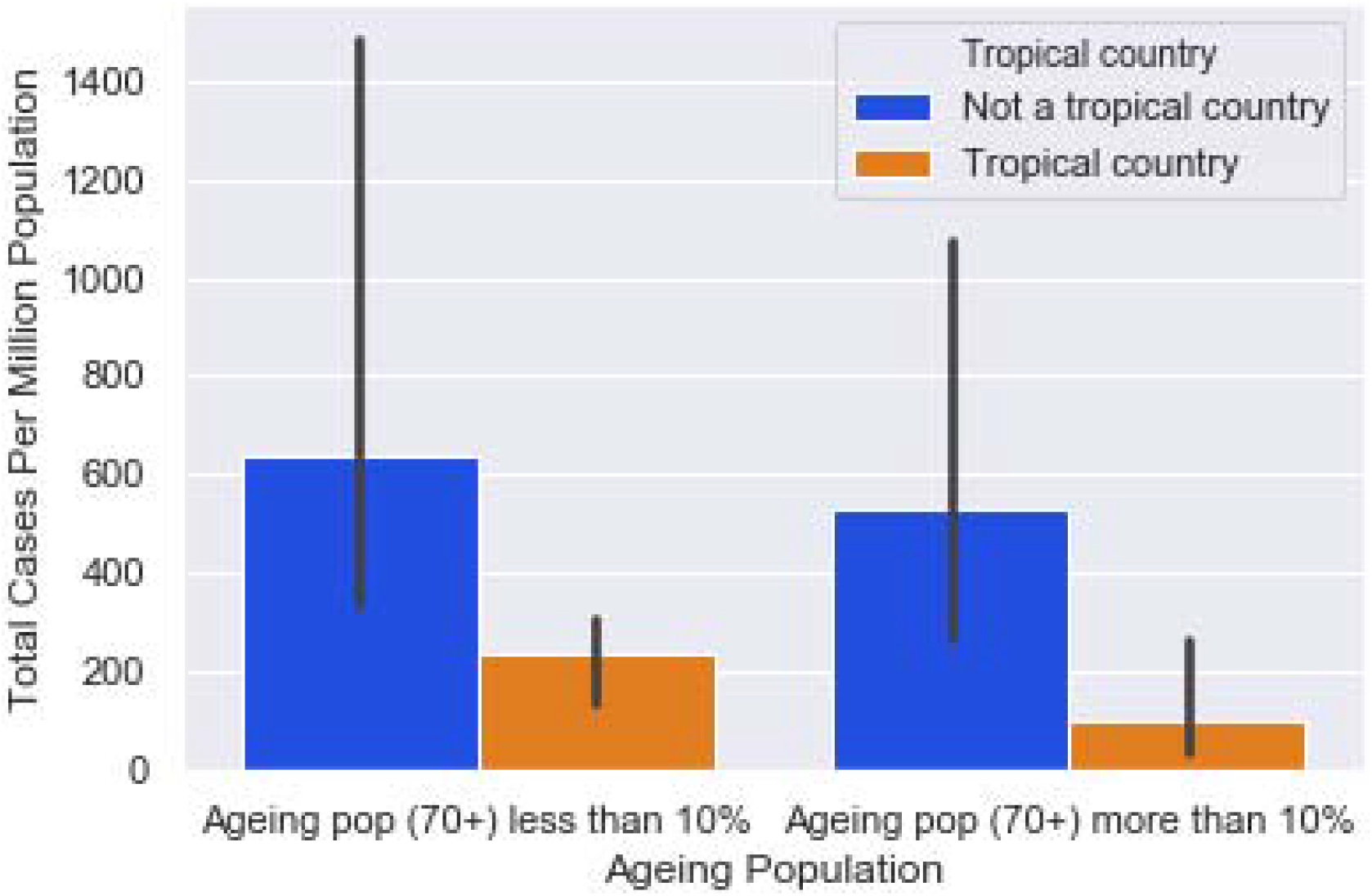
Case fatality rate by age

**Figure 6:**
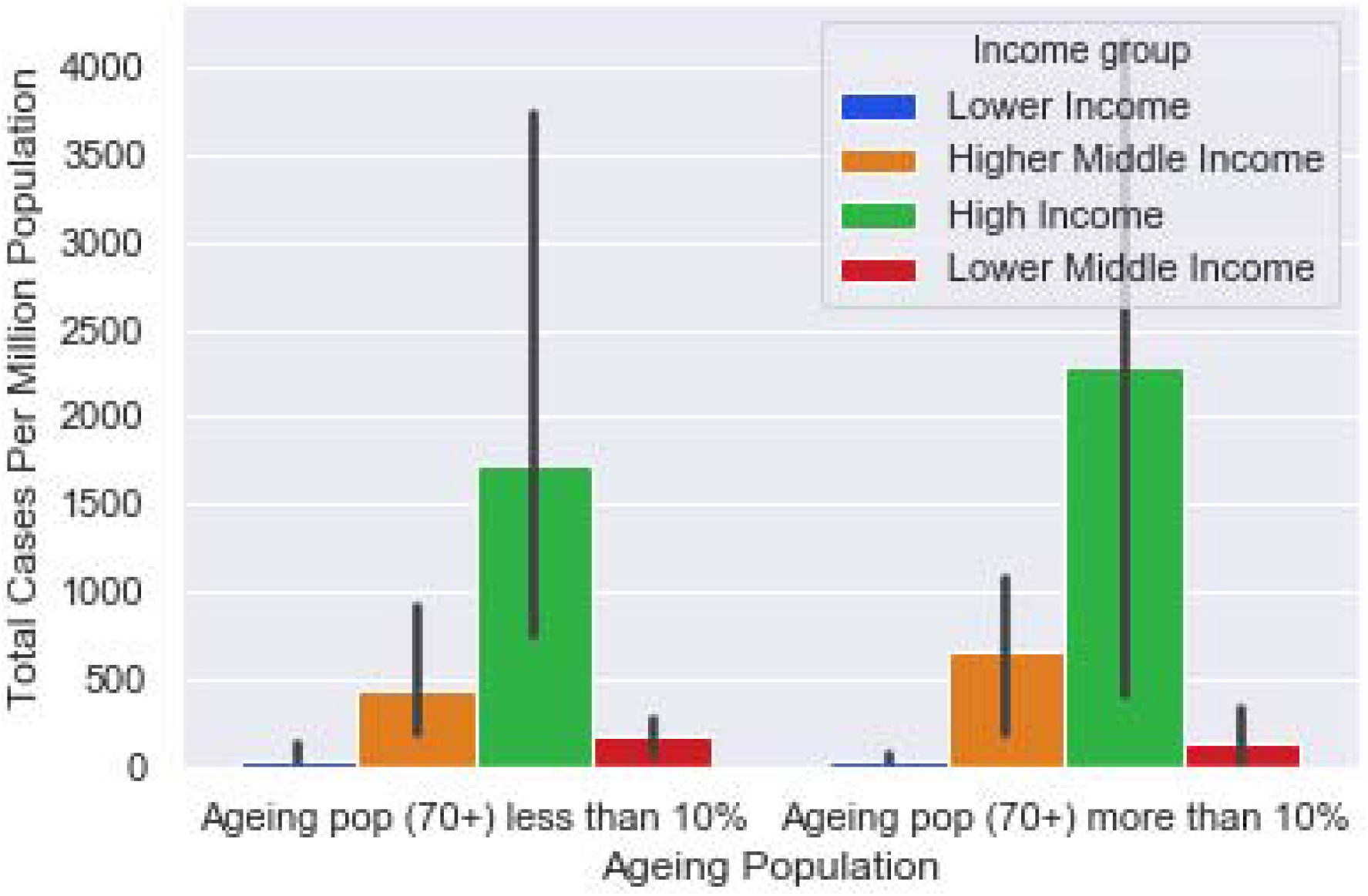
Total infection by country type and ageing population

The average case fatality rate (CFR) is 3.55 with a maximum of 21.99. Average tests per thousand and average cases per million are 39 and 1,471, respectively, with a maximum of 241 and 26,583. About 6% of the population ages 70 years and above and 39% countries are located in tropics. However, there is a large variation in the proportion of ageing population ranging from .53 to 18.49. So far evidence suggests that ageing population is more vulnerable to COVID-19 and therefore the variation in ageing population across countries can significantly predict the death rates of the countries.

Average population density per square kilometer is 265, with a maximum of 7,915. Countries have average of 60% urban population, whereas seven countries including Bermuda, Gibraltar, Hong Kong, Kuwait, Singapore, Cayman Islands, and Sint Maarten (Dutch part) have 100% of its population living in urban areas. Average cardiovascular diseases (CVD) death rate was 256 and there were 3 beds per 1,000 population in the countries. Japan has the highest (13.63) hospital beds per 1,000 population. About 88% children ages 12-23 months are immunized and Ethiopia has the lowest rate (41.18%). Montenegro has the highest smoking prevalence rate of 47.03% whereas the average rate is 22%. Besides, average extreme poverty rate was 14% with a maximum of 77.6% in Madagascar.

Table 2 reports the findings from *t*-test of several variables on case fatality rate and total cases per million. Probabilities [Pr (T < t)] are reported here. Notably, we do not find a statistically significant difference in the means of aging population (70 years and above), hospital bed per 1,000 population, smoking prevalence. Meanwhile, we notice that Urban population (p=0.0000), tropical country (p=0.0053), CVD death rate (p=0.0478), and immunization (p=0.0078) have a significant difference in their means.

**Table 2:** Findings from t-test.

|  | CFR |  |  | Total Cases Per Million |  |  |
| --- | --- | --- | --- | --- | --- | --- |
| | $H_0: \text{diff}=0$ | | | $H_0: \text{diff}=0$ | | |
| | $H_A: \text{diff} < 0$ | $H_A: \text{diff} \neq 0$ | $H_A: \text{diff} > 0$ | $H_A: \text{diff} < 0$ | $H_A: \text{diff} \neq 0$ | $H_A: \text{diff} > 0$ |
| Ageing population | 0.1490 | 0.2980 | 0.8510 | 0.9108 | 0.1785 | 0.0892 |
| Hospital bed per 1,000 population | 0.2043 | 0.4085 | 0.7957 | 0.8499 | 0.3001 | 0.1501 |
| Urban population | 0.1046 | 0.2091 | 0.8954 | 0.0000 | 0.0000 | 1.0000 |
| Topical country | 0.7187 | 0.5626 | 0.2813 | 0.9974 | 0.0053 | 0.0026 |
| CVD death rate | 0.8946 | 0.2109 | 0.1054 | 0.9761 | 0.0478 | 0.0239 |
| Immunization | 0.2057 | 0.4114 | 0.7943 | 0.0039 | 0.0078 | 0.9961 |
| Smoking prevalence | 0.1859 | 0.3718 | 0.8141 | 0.5752 | 0.8495 | 0.4248 |

Table 3 shows pairwise correlation matrix of the variables. Results show that aging population is positively correlated with CFR and CVD death rate have significant negative correlation with case fatality rate (CFR). On the contrary, population density, urban population, and immunization are positively correlated with total cases per million, and tropical country, CVD death rate, and extreme poverty rate have significant negative correlation with total cases per million. However, all the significant correlations are at a significance level of 10%.

**Table 3:**
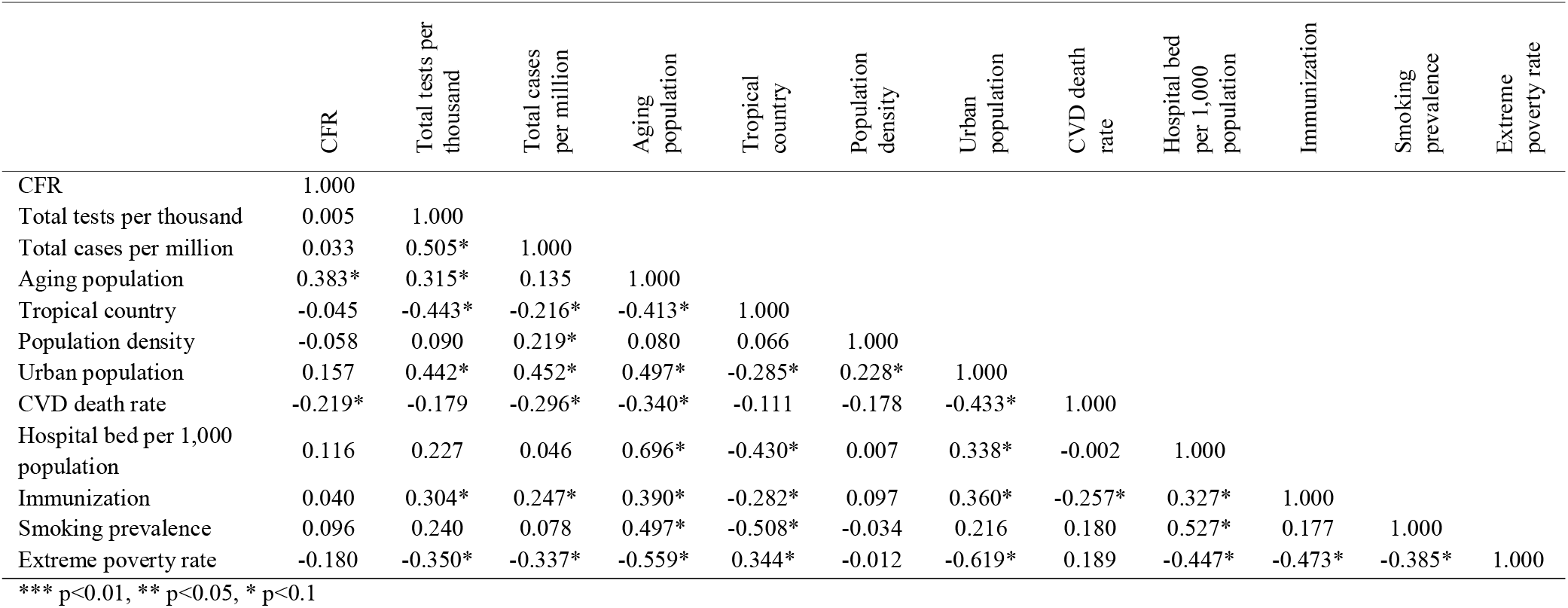
Pairwise correlation matrix.

|  | CFR | Total tests per thousand | Total cases per million | Aging population | Tropical country | Population density | Urban population | CVD death rate | Hospital bed per 1,000 population | Immunization | Smoking prevalence | Extreme poverty rate |
| --- | --- | --- | --- | --- | --- | --- | --- | --- | --- | --- | --- | --- |
| CFR | 1.000 |  |  |  |  |  |  |  |  |  |  |  |
| Total tests per thousand | 0.005 | 1.000 |  |  |  |  |  |  |  |  |  |  |
| Total cases per million | 0.033 | 0.505* | 1.000 |  |  |  |  |  |  |  |  |  |
| Aging population | 0.383* | 0.315* | 0.135 | 1.000 |  |  |  |  |  |  |  |  |
| Tropical country | -0.045 | -0.443* | -0.216* | -0.413* | 1.000 |  |  |  |  |  |  |  |
| Population density | -0.058 | 0.090 | 0.219* | 0.080 | 0.066 | 1.000 |  |  |  |  |  |  |
| Urban population | 0.157 | 0.442* | 0.452* | 0.497* | -0.285* | 0.228* | 1.000 |  |  |  |  |  |
| CVD death rate | -0.219* | -0.179 | -0.296* | -0.340* | -0.111 | -0.178 | -0.433* | 1.000 |  |  |  |  |
| Hospital bed per 1,000 population | 0.116 | 0.227 | 0.046 | 0.696* | -0.430* | 0.007 | 0.338* | -0.002 | 1.000 |  |  |  |
| Immunization | 0.040 | 0.304* | 0.247* | 0.390* | -0.282* | 0.097 | 0.360* | -0.257* | 0.327* | 1.000 |  |  |
| Smoking prevalence | 0.096 | 0.240 | 0.078 | 0.497* | -0.508* | -0.034 | 0.216 | 0.180 | 0.527* | 0.177 | 1.000 |  |
| Extreme poverty rate | -0.180 | -0.350* | -0.337* | -0.559* | 0.344* | -0.012 | -0.619* | 0.189 | -0.447* | -0.473* | -0.385* | 1.000 |

\*\*\* $p < 0.01$ , \*\* $p < 0.05$ , \* $p < 0.1$

From the *t*-test and correlational analysis we find some expected results such as a positive correlation between ageing and death rate; a positive correlation between the proportion of urban population and infection; negative correlation of tropical country status with both infections and death rates and other interesting results have emerged. However, *t*-test and correlation cannot take other controls/confounding variables in account; and therefore, a meaningful association cannot be observed. The regression can help better understand the association. It is worth mentioning, even though regression is mostly used for understanding causation, regression cannot show the causation with a clear identification strategy such as randomized experiment. Otherwise, regression will show most the correlation.

Table 4 presents the regression estimates. First two column shows the OLS and LASSO estimates for case fatality rate and the third column shows OLS estimates for total cases per million.

**Table 4:** Regression estimates.

|  | (1)<br>CFR_OLS | (2)<br>CFR_LASSO | (3)<br>Total Cases Per Million |
| --- | --- | --- | --- |
| Population ages 70 years and above (%<br>of total population) | 0.586***<br>(0.124) | 0.705***<br>(0.184) | -49.147<br>(96.336) |
| Tropical country dummy | -0.041<br>(1.026) | 0.303<br>(0.829) | -231.955<br>(680.691) |
| Population density | -0.000<br>(0.000) | -0.001*<br>(0.000) | -0.406<br>(1.269) |
| Urban Population<br>(% of total population) | 0.027<br>(0.023) | 0.035<br>(0.028) | 34.324*<br>(18.187) |
| CVD death Rate | -0.002<br>(0.004) | 0.001<br>(0.003) | -5.655*<br>(2.897) |
| Hospital bed per 1,000 population | -0.435**<br>(0.191) | -0.500***<br>(0.185) | -146.355<br>(135.223) |
| Total tests per thousand | -0.016*<br>(0.009) | -0.018*<br>(0.010) | 19.264***<br>(6.979) |
| Immunization, measles<br>(% of children ages 12-23 months) | -0.025<br>(0.060) | -0.017<br>(0.041) | -26.514<br>(36.325) |
| Smoking prevalence, total (ages 15+) |  |  | 28.026<br>(35.725) |
| Extreme poverty rate |  |  | -0.855<br>(36.072) |
| Constant | 2.644<br>(5.538) |  | 2906.861<br>(3673.821) |
| Observations<sup>†</sup> | 78 | 75 | 60 |
| Adjusted $R^2$ | 0.320 | | 0.359 |

Standard errors in parentheses
\* $p < .10$ , \*\* $p < .05$ , \*\*\* $p < .01$
<sup>†</sup> Number of observation varies from descriptive statistics based on the availability of data for observations

From the OLS estimates we see that aging population and hospital bed per 1,000 population have significant effect on case fatality rate (p<0.01). CFR increases by 0.586 with aging population increased by 1% of the total population. It also reveals that one unit increase in hospital bed per 1,000 population results in 0.435 unit reduction in CFR. For the LASSO estimates, consistent results are detected for both of the estimates (p<0.01). Coefficients are 0.705 and −0.500 for aging population and hospital bed per 1,000 population.

On another note, total cases per million is determined by total tests per thousand. Apparently, it features that one unit increment in total tests per thousand results in an increase of 19.26 units in total cases per million (p<0.01). In particular, it symbolizes the more a country tests COVID-19, the more cases found. At the same time, 1% increase in urban population significantly (p<0.10) results in an increase of 34.32 cases per million. Most other covariates are found to be less relevant (insignificant) in explaining infection and death rates.

Based the cross-sectional analysis of COVID-19 cases, we observed that demographic characteristics and country’s health systems capacity have strong correlation with COVID-19 deaths. On the other hand, a very few predictors of infection can be obtained. However, some key findings have been observed from this analysis.

First, the country with larger number of ageing population had to face higher death rates. It is not surprising, Various studies found that ageing population is most vulnerable, and so countries with higher proportion of aging population has higher death rates.

Second, a proxy of health system variable—hospital beds per capita—appears to be highly important in COVID-19. It is not surprising since more hospital beds means the patients will not overwhelm the health systems, and thus treatment can be ensured for many and therefore, people will not die just due to lack of basic treatment say oxygen. In addition, more hospitals are also likely come with more ICU beds which again can save more lives.

Third, whenever it comes to infection, story seems to be still unknown; the puzzle still remains. However, we find the urbanization has a strong correlation with infection. It is surprising since urban areas are highly densed and the individuals may have higher interactions and transmissions. Most of worst affected countries also had worst affected cities such as New York, London.

## 5. Conclusion

Using cross country data, the study attempted to understand the variation of case fatality rates and infection rate of COVID-19 across countries. The study finds that countries health systems and demographic structures play a significant role in determining death rates. The countries with higher aging population will have higher death rates, and country with good health system will have lower death rates. In addition, countries with higher urbanization would experience more severe outbreak. Other variables such as weather, vaccination coverage, etc. do not have association with neither death rates nor level of infections. The study suggests to improve the system at a maximum capacity to reduce the death rates, while special attention in social distancing should be enforced for the entire country, for that matter, highly-dense urban areas. Since pandemic is still on rise in many countries in this conclusion may change in the long run. Future research especially when pandemic will be almost over, can address those issues.

## Data Availability

The study uses publicly available data that has been cited with the proper links. However, authors are ready to share data if requested.

## Appendix

## Footnotes

1 https://news.google.com/covid19/map?hl=en-US&gl=US&ceid=US%3Aen

2 https://ourworldindata.org/coronavirus-data

3 https://databank.worldbank.org/source/world-development-indicators#

4 https://worldpopulationreview.com/countries/tropical-countries/?fbclid=IwAR3lkkH3tseEu3WB7aIZBYjwAXj_ERrpbGHzTTl8MtjiymK0lG8y4dDL6lQ

